# Biomarkers of atrial fibrillation-related pathways and left atrial structure and function in an overweight and obese population

**DOI:** 10.1101/2024.09.17.24313430

**Authors:** Linzi Li, Dora Romaguera, Angel M. Alonso-Gómez, Estefanía Toledo, Amit J. Shah, Marta Noris Mora, Lucas Tojal-Sierra, Miguel Angel Martinez-Gonzalez, Caterina Mas-Llado, Cristina Razquin, Ramón Estruch, Montserrat Fitó, Alvaro Alonso

**Author notes:** Corresponding author: Linzi Li, MPH, MSPH.

## Abstract

**Background:** Exploring longitudinal associations of blood biomarkers with left atrial (LA) structure and function can enhance our understanding of atrial fibrillation (AF) etiopathogenesis.

**Methods:** We studied 532 participants of the PREDIMED-Plus trial, a multicenter randomized trial in overweight and obese adults with metabolic syndrome. At baseline, 3 and 5 years after randomization, participants underwent transthoracic echocardiography and provided blood for serum biomarker measurements [propeptide of procollagen type I (PICP), high-sensitivity (hs) troponin T (hsTnT), hs C-reactive protein (hsCRP), 3-nitrotyrosine (3-NT), and N-terminal propeptide of B-type natriuretic peptide (NT-proBNP)]. Outcomes of interest included LA peak systolic longitudinal strain (LA PSLS), LA volume index (LAVi), LA function index (LAFi), and LA stiffness index (LASi). We performed cross-sectional and longitudinal analyses to evaluate relationships between log-transformed biomarkers and echocardiographic measurements using multiple linear regression and mixed models.

**Results:** The participants in this analysis had a mean age of 65.0 (SD 4.8) years, and 40% were females. At baseline, increased NT-proBNP and hsTnT were associated with larger LAVi and worse LA function as measured by the LAFi, LASi, and LA PSLS. Longitudinally, higher NT-proBNP, but not higher hsTnT, was associated with increased LAVi and worsening LA function. Over 5 years, 1 unit increase in log(NT-proBNP) was associated with steeper decline in LA PSLS (-0.19%, 95% CI -0.35%, -0.02%) and greater increase in LAVi (0.28 mL/m2, 95% CI 0.10, 0.45) each year. PICP, hsCRP, and 3-NT did not show consistently significant associations with LA outcomes at baseline and through 5 years.

**Conclusion:** In an overweight and obese population, higher NT-proBNP was associated with LA volume enlargement and worsening LA function over 5 years. The implications of these findings for the prevention and prediction of AF warrant further investigation.

## Introduction

Atrial fibrillation (AF), a common cardiac arrhythmia, is a risk factor of many adverse outcomes, including stroke, heart failure, and death.^1^ It has been estimated that by 2050, 5.6 million people will be living with AF in the United States.^2^ While AF results in a huge disease burden on the population, effective detection and prevention strategies are still lacking due to the incomplete understanding of AF etiopathogenesis. In animal models and AF patients, researchers have identified evidence indicating that particular electrical triggers producing an effect on a vulnerable structural and functional atrial substrate may play a role in AF onset.^3^ In addition, previous work has suggested that atrial remodeling and fibrosis are significant features of the substrate facilitating AF development, which involves multifactorial processes and various mechanisms.^4^ Atrial remodeling and fibrosis can be attributed to well-known AF risk factors, such as obesity and hypertension.^5,6^ In addition, circulating biomarkers of atrial fibrosis,^7^ myocardial injury,^8^ and atrial stretch and overload ^9^ were elevated among individuals with higher AF risk.

Extensive research has focused on the underlying mechanisms of abnormal AF substrate and how risk factors contribute to it. Echocardiography of the left atrium is a non-invasive and capable approach to assess the atrial substrate (remodeling and fibrosis). It has been found that larger left atrial (LA) size, volume and function, markers of atrial structural remodeling, are risk factors of AF.^10–12^ Furthermore, known AF pathway-related blood biomarkers can characterize the atrial substrate facilitating AF onset. Previous studies have found several blood biomarkers of AF-related pathways associated with increased risk of AF: carboxy-terminal propeptide of procollagen type I (PICP, marker of cardiac fibrosis), high-sensitivity troponin T (hsTnT, marker of myocardial damage), high-sensitivity C reactive protein (hsCRP, marker of inflammation), 3-nitrotyrosine (3-NT, marker of oxidative stress), and N-terminal propeptide of B-type natriuretic peptide (NT-proBNP, marker of atrial stretch).^13–18^ However, there is little information regarding how blood biomarkers are associated with LA structure and function. A better understanding of the initiation and development of AF substrate would help identify effective AF prevention strategies. Therefore, we propose investigating the association between biomarkers of AF-related pathways and LA structure and function in an obese and overweight population with metabolic syndrome that is at higher risk of AF.

## Methods

### Study design and population

This study was performed in the PREDIMED-Plus study, a multicenter randomized trial in overweight and obese adults focusing on the primary prevention of cardiovascular disease (CVD).^19^ From 2013 to 2016, 6874 individuals (3,574 men aged 55-75 and 3,300 women aged 60-75) who met at least three components of the metabolic syndrome and had a body mass index (BMI) ≥ 27 and < 40 kg/m^2^ were recruited from 23 study centers in Spain and randomized 1:1 to an intensive lifestyle intervention (ILI) program or a control group.^19^ ILI program was based on an energy-reduced Mediterranean diet (erMedDiet), increased physical activity, and cognitive-behavior weight support, and the control intervention was a low-intensity dietary advice on the Mediterranean diet which was not energy-restricted. The intervention period spanned 6 years, concluding at the end of 2022. Participants are being followed up annually over an 8-year period, which is scheduled to conclude at the end of 2024. Baseline and follow-up examinations have been performed at baseline, at 6 months and year 1 post-randomization, and annually thereafter. As part of an ancillary study to the PREDIMED-Plus trial, echocardiographic studies were conducted and blood biomarkers were measured in a sub-sample from 3 PREDIMED-Plus study sites (University of Navarra, Araba University Hospital, Son Espases University Hospital; n = 566) at baseline and years 3 and 5 after randomization.^20^ The trial was registered in 2014 at [www.isrctn.com/ISRCTN89898870]. The full protocol is available at https://www.predimedplus.com/en/project/.

In this analysis, we included participants who completed the baseline exam and excluded those who had AF, all the blood biomarker measurements and LA structure and function measurements missing at baseline. Five hundred and thirty-two subjects were kept in the analyzed dataset. For the longitudinal analysis in this study, we also excluded participants who 1) did not complete any follow-up exam; 2) lacked all biomarkers and LA structure and function measurements at year 3 and year 5 exams (n=529). For models that examined the effect of changes in biomarker concentrations between baseline and years 3 and 5, those who had evidence of AF at year 3 or year 5 exams were excluded, respectively. Figure 1 shows the CONSORT chart of this study.

**Figure 1.**
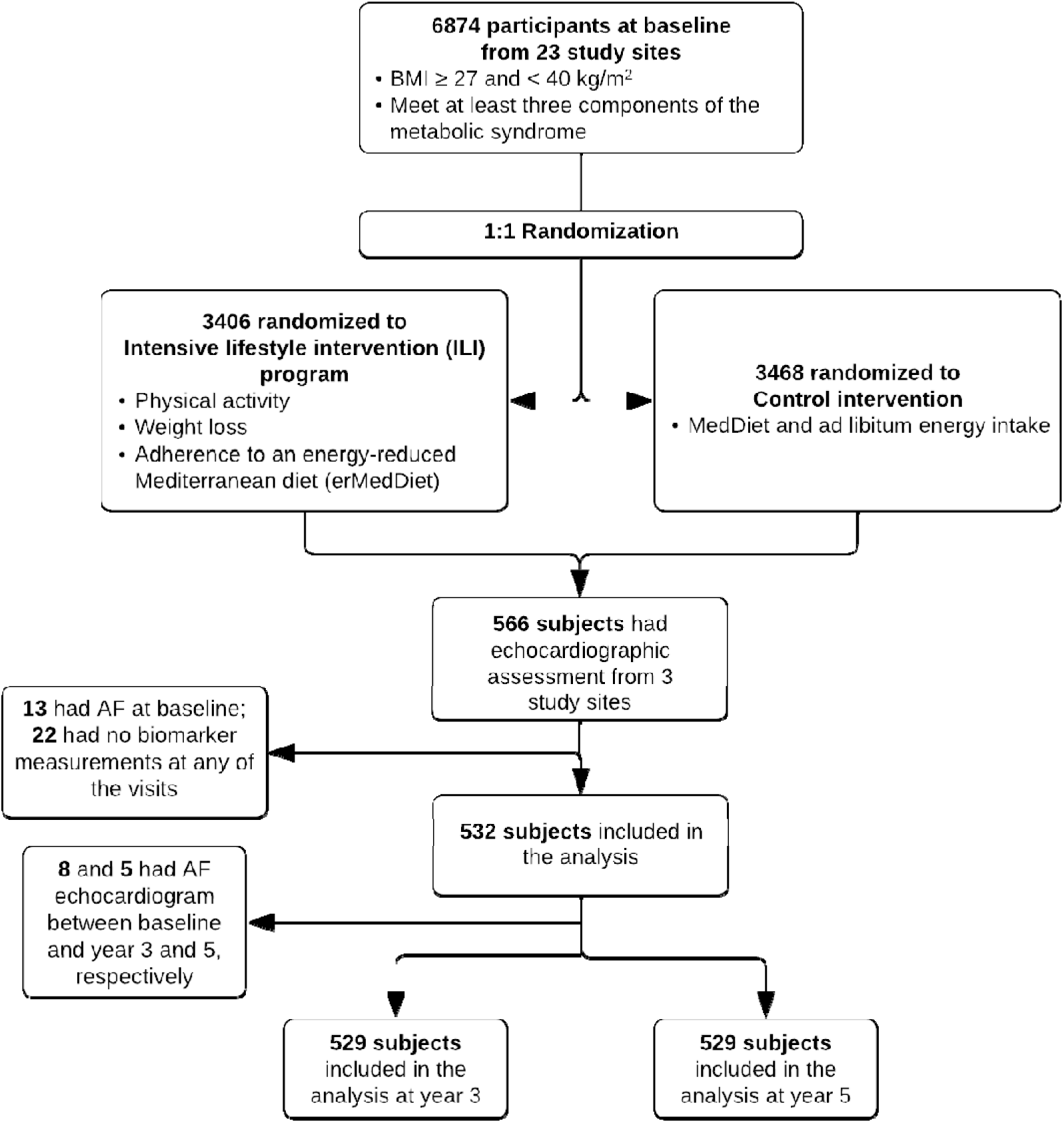
CONSORT chart of the study sample, PREDIMED-Plus trial *The count of exclusion numbers may not necessarily match the count of subjects removed due to overlap.

### Blood biomarkers and LA structure and function

The exposures were concentrations of five AF-related blood biomarkers: PICP (fibrosis), hsTnT (myocardial damage), hsCRP (inflammation), 3-NT (oxidative stress), and NT-proBNP (atrial stretch). They were measured from serum samples collected at baseline and two follow-up exams at years 3 and 5. 3-NT and PICP were measured by commercially available enzyme-linked immunosorbent assay technique (ELISA). Human Nitrotyrosin ELISA kit (Abcam, Cambridge, UK) and MicroVue PICP EIA (Quidel, San Diego, CA, USA) were used for 3-NT and PICP, respectively. HsCRP was measured by immunoturbidimetry on a Cobas 8000 autoanalyzer (Roche Diagnostics); hsTnT and NT-proBNP were measured by electrochemiluminescence immunoassay (ECLIA). All the biomarker analyses were performed blindly.

The primary outcomes of this study were LA function and structure measures, including LA conduit strain (LAScd), LA contractile strain (LASct), LA peak systolic longitudinal strain (LA PSLS), max LA volume (LAVmax), LA volume index (LAVi), total LA ejection fraction (LAEF), LA function index (LAFi), and LA stiffness index (LASi). At baseline, years 3 and 5 exams, participants at the 3 sites underwent transthoracic echocardiography (TTE) to obtain their LA outcome measures. Standard TTE was performed using Vivid 7 or Vivid 9 (GE Healthcare) machines at each site following the procedures described elsewhere.^21,22^ All echocardiographic studies were transferred to a core laboratory and blindly read by two cardiologists. All assessments for a given measure were done by the same cardiologist. The calculations for the derived measures were as follows:

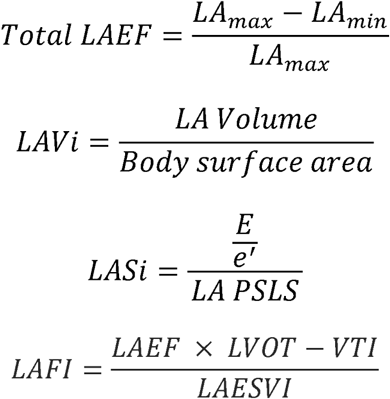

LAEFI: maximal left atrial volume in end systole indexed to body surface area

LVOT: left ventricular outflow tract

VTI: velocity time integral

LAESVI: left atrial end-systolic volume index

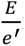: early diastolic mitral annular flow velocity/mitral annulus early diastolic velocity

### Other covariates

The covariates assessed at baseline considered in the analyses include demographic characteristics (age, sex, national origin), behavioral characteristics (smoking status, alcohol consumption), clinical measurements (BMI, systolic blood pressure [SBP], diastolic blood pressure [DBP], low-density lipoprotein cholesterol [LDLc], high-density lipoprotein cholesterol [HDLc], estimated glomerular filtration rate [eGFR]), medications (anti-hypertension, lipid-lowing medication), history of diabetes, and intervention group.

### Statistical Methods

Biomarker concentrations were log-transformed and these values were used in all analyses. Left atrial strain outcomes (LAScd and LASct) are presented and modeled as absolute values so that a lower value implies a worse function throughout. Multiple linear regression models were employed to examine the cross-sectional association between biomarker concentrations and LA outcomes. Then mixed models were used to estimate: 1) the effect of biomarker concentrations at baseline on changes in LA outcomes between baseline and year 5, 2) the effect of changes in biomarker concentrations on concurrent changes in LA outcomes over 5 years of follow-up, and 3) the effect of changes in biomarker concentrations between baseline and year 3 on LA outcome changes between year 3 and year 5. The changes in biomarker concentrations were calculated as log(changes in biomarker concentration) = log(biomarker concentration at year 3 or 5) – log(biomarker concentration at baseline). Random effects for the individual and family unit were included in the mixed models because 24 subjects had a single household cohabitant participating in the study. The time of follow-up was modeled as a categorical variable (year 3 vs. baseline and year 5 vs. baseline). Both the cross-sectional and longitudinal analyses deployed the following adjustments: 1) adjusted for age and sex; 2) further adjusted for BMI, ethnic origin, smoking status, alcohol consumption, SBP, DBP, HDLc, LDLc, anti-hypertension medication, lipid-lowing medication, history of diabetes, and eGFR. In the abovementioned longitudinal analyses 2) and 3), the trial intervention group was also adjusted for. The results were presented as the coefficients of the models, along with 95% confidence intervals (CIs). All the analyses were performed with SAS 9.4 (Cary, NC; SAS Institute Inc.).

## Results

In this study, 532 subjects were included, with a mean age of 65 years (standard deviation [SD] 4.9) and a mean BMI of 32.2 kg/m^2^ (SD 3.3). Additionally, 40% (214) of the participants were female. At baseline, the mean LAScd was 12.0% (SD 4.4), mean LASct 15.7% (SD 4.6), and mean LA PSLS 27.6% (SD 6.5). The mean LAEF was 59% (SD 9.3) and mean LAVmax 44 mL (SD 14.4). The mean LAVi was 22.7 (SD 7.0), mean LAFi 67.9 (SD 29.4), and mean LASi 0.4 (SD 0.2) (Table 1). Throughout the follow-up, LAScd, LASct, LAVmax, LAVi, and LASi increased, while LA PSLS, LAEF, and LAFi decreased. (Figure 2)

**Figure 2.**
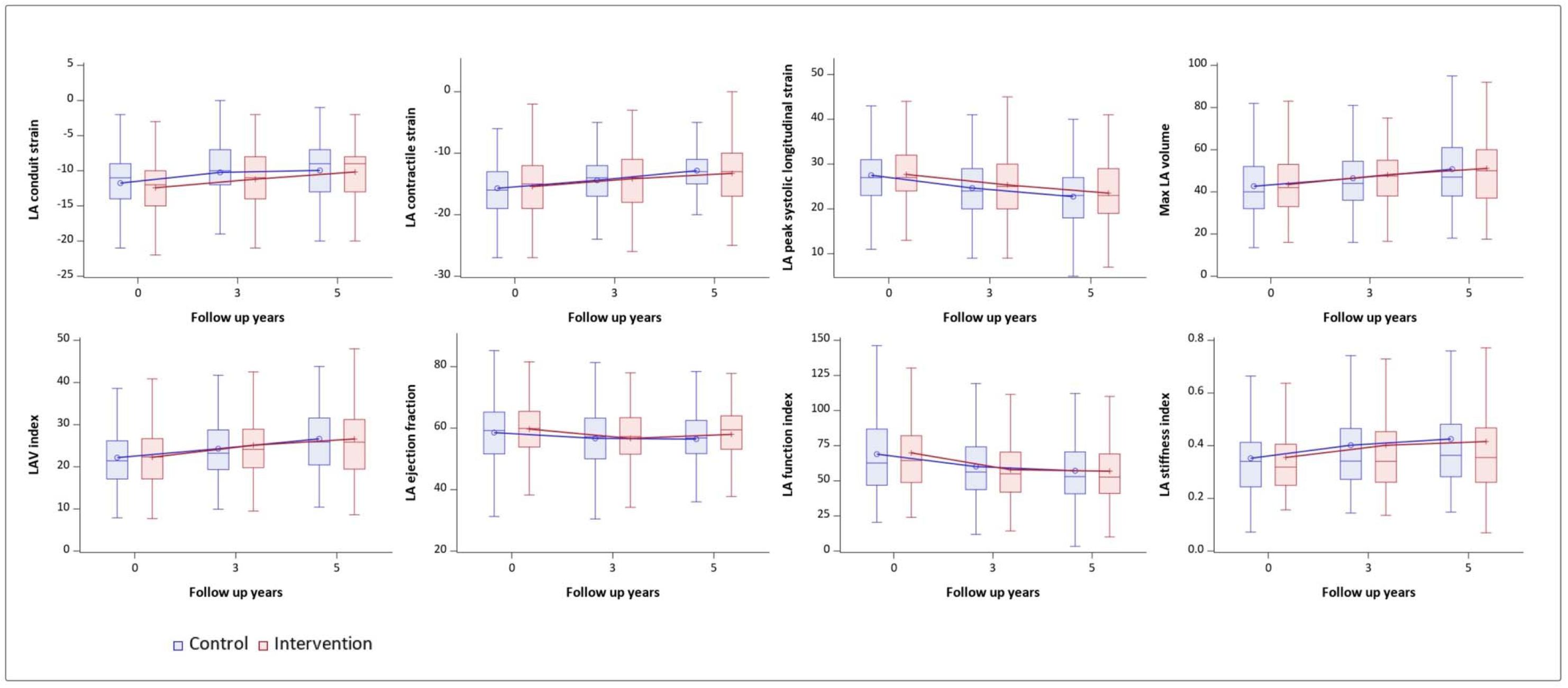
Changes of LA outcomes at baseline, years 3 and 5 TTE exams

**Table 1.**
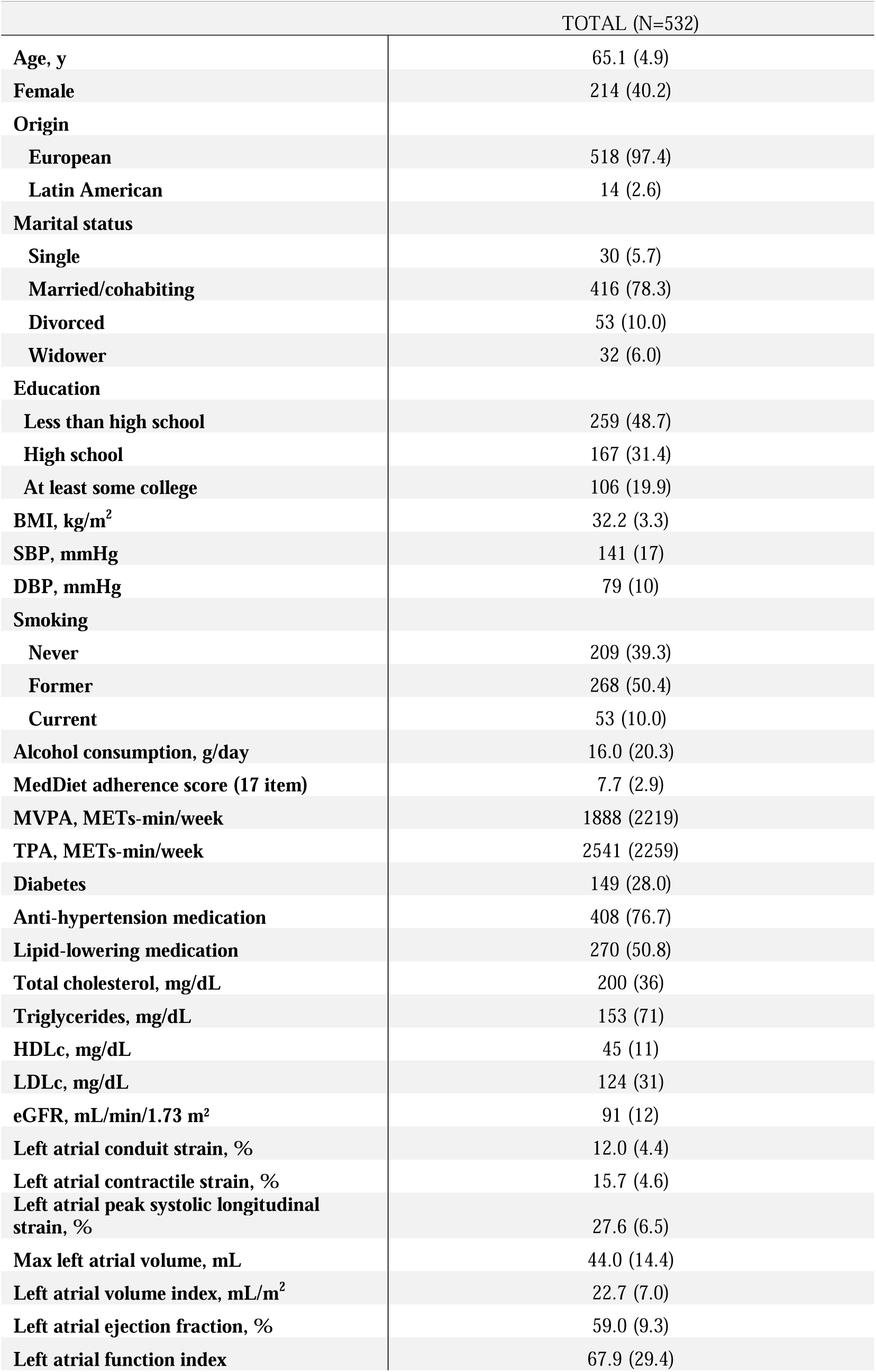
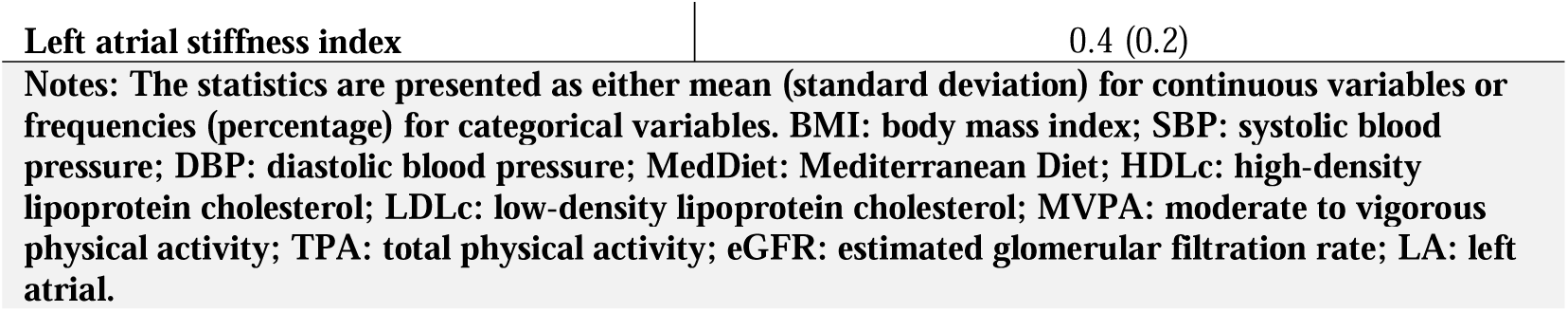
Baseline characteristics of participants in the PREDIMED-Plus trial included in this study.

At baseline, a higher hs-cTnT concentration was associated with a lower LA PSLS (-1.54, 95% CI: -3.03, -0.06), larger LAVmax (3.44, 95% CI: 0.23, 6.66), and worse LAFi (-6.83, 95% CI: -13.64, -0.02) and LASi (0.04, 95% CI: 0.01, 0.08), after adjusting for all the covariates. An elevated NT-proBNP was found to be linked with decreases in LASct (-0.56, 95% CI -1.07, -0.05), larger LAVmax (1.84, 95% CI 0.36, 3.32) and LAVi (0.78, 95% CI: 0.04, 1.52), and worse LAFi (-3.61, 95% CI: 6.75, -0.46) and LASi (0.02, 95% CI: 0.00, 0.04), after adjusting for all the covariates. No consistent associations were found between PICP, CRP, and 3-NT with LA outcomes when considering all the covariates (Table 2).

**Table 2.** Associations between atrial fibrillation-related biomarkers and left atrial function and structure at baseline, PREDIMED-Plus trial.

|  |  | PICP | hs-cTnT | CRP | 3-NT | NT-proBNP |
| --- | --- | --- | --- | --- | --- | --- |
| **Left atrial conduit strain, %** |  |  |  |  |  |  |
|  | Crude | -0.03 (-0.93, 0.87) | -0.96 (-1.84, -0.08) | 0.31 (-0.10, 0.71) | 0.32 (-0.10, 0.74) | -0.08 (-0.52, 0.37) |
|  | Model 1 | -0.13 (-1.02, 0.76) | -1.03 (-1.99, -0.08) | 0.30 (-0.10, 0.70) | 0.34 (-0.08, 0.76) | 0.22 (-0.25, 0.68) |
|  | Model 2 | -0.05 (-0.95, 0.84) | -0.71 (-1.75, 0.33) | 0.16 (-0.27, 0.59) | 0.34 (-0.09, 0.77) | 0.18 (-0.30, 0.66) |
| **Left atrial contractile strain, %** |  |  |  |  |  |  |
|  | Crude | 0.20 (-0.74, 1.15) | -0.69 (-1.62, 0.24) | 0.22 (-0.21, 0.64) | -0.19 (-0.63, 0.26) | -0.86 (-1.32, -0.40) |
|  | Model 1 | 0.17 (-0.77, 1.11) | -1.25 (-2.27, -0.24) | 0.27 (-0.15, 0.69) | -0.16 (-0.60, 0.29) | -0.75 (-1.23, -0.26) |
|  | Model 2 | 0.24 (-0.71, 1.19) | -0.90 (-2.01, 0.20) | 0.28 (-0.18, 0.73) | -0.17 (-0.63, 0.29) | -0.56 (-1.07, -0.05) |
| **Left atrial peak systolic longitudinal strain, %** |  |  |  |  |  |  |
|  | Crude | 0.11 (-1.19, 1.42) | -1.76 (-3.05, -0.48) | 0.51 (-0.08, 1.10) | 0.15 (-0.47, 0.77) | -0.87 (-1.52, -0.22) |
|  | Model 1 | -0.03 (-1.31, 1.26) | -2.26 (-3.65, -0.87) | 0.53 (-0.05, 1.12) | 0.21 (-0.40, 0.82) | -0.47 (-1.14, 0.20) |
|  | Model 2 | 0.14 (-1.13, 1.41) | -1.54 (-3.03, -0.06) | 0.40 (-0.21, 1.02) | 0.19 (-0.44, 0.81) | -0.31 (-1.00, 0.38) |
| **Max left atrial volume, mL** |  |  |  |  |  |  |
|  | Crude | 2.22 (-0.65, 5.10) | 5.85 (3.04, 8.67) | -1.17 (-2.47, 0.13) | -0.70 (-2.08, 0.67) | 1.46 (0.02, 2.90) |
|  | Model 1 | 2.32 (-0.50, 5.13) | 3.92 (0.85, 6.98) | -0.83 (-2.11, 0.46) | -0.48 (-1.83, 0.86) | 2.24 (0.77, 3.71) |
|  | Model 2 | 1.89 (-0.85, 4.63) | 3.44 (0.23, 6.66) | -1.05 (-2.39, 0.28) | -0.27 (-1.61, 1.07) | 1.84 (0.36, 3.32) |
| **Left atrial volume index, mL/m<sup>2</sup>** |  |  |  |  |  |  |
|  | Crude | 0.85 (-0.55, 2.26) | 1.79 (0.40, 3.18) | -0.74 (-1.37, -0.10) | -0.18 (-0.85, 0.50) | 1.07 (0.37, 1.77) |
|  | Model 1 | 0.95 (-0.45, 2.36) | 1.60 (0.07, 3.13) | -0.69 (-1.33, -0.05) | -0.17 (-0.84, 0.51) | 1.03 (0.29, 1.76) |
|  | Model 2 | 0.78 (-0.60, 2.15) | 1.58 (-0.03, 3.19) | -0.58 (-1.25, 0.08) | -0.12 (-0.79, 0.55) | 0.78 (0.04, 1.52) |
| **Left atrial ejection fraction, %** |  |  |  |  |  |  |
|  | Crude | -0.58 (-2.44, 1.29) | -0.24 (-2.10, 1.61) | 0.35 (-0.50, 1.20) | 0.31 (-0.58, 1.20) | -1.79 (-2.71, -0.86) |
|  | Model 1 | -0.63 (-2.48, 1.21) | -1.57 (-3.57, 0.44) | 0.50 (-0.34, 1.34) | 0.43 (-0.45, 1.31) | -1.43 (-2.39, -0.47) |
|  | Model 2 | -0.52 (-2.37, 1.34) | -2.21 (-4.38, -0.04) | 0.30 (-0.60, 1.20) | 0.39 (-0.52, 1.29) | -1.53 (-2.53, -0.54) |
| **Left atrial function index** |  |  |  |  |  |  |
|  | Crude | -3.78 (-9.75, 2.19) | -6.01 (-11.87, -0.15) | 2.00 (-0.70, 4.69) | 0.80 (-2.03, 3.62) | -4.11 (-7.07, -1.15) |
|  | Model 1 | -4.00 (-9.99, 1.98) | -6.69 (-13.16, -0.23) | 1.95 (-0.77, 4.67) | 0.83 (-2.00, 3.67) | -4.03 (-7.14, -0.91) |
|  | Model 2 | -2.87 (-8.73, 2.99) | -6.83 (-13.64, -0.02) | 0.65 (-2.18, 3.49) | 0.96 (-1.87, 3.80) | -3.61 (-6.75, -0.46) |
| **Left atrial stiffness index** |  |  |  |  |  |  |
|  | Crude | -0.01 (-0.04, 0.03) | 0.04 (0.01, 0.07) | 0.00 (-0.02, 0.01) | 0.01 (-0.01, 0.03) | 0.04 (0.03, 0.06) |
|  | Model 1 | 0.00 (-0.03, 0.03) | 0.06 (0.02, 0.09) | 0.00 (-0.02, 0.01) | 0.01 (-0.01, 0.02) | 0.03 (0.01, 0.05) |
|  | Model 2 | -0.01 (-0.04, 0.03) | 0.04 (0.01, 0.08) | 0.00 (-0.02, 0.01) | 0.01 (-0.01, 0.02) | 0.02 (0.00, 0.04) |

**Notes:** Mixed models were used. Model 1: adjusted for age, sex; Model 2: further adjusted for further adjusted for BMI, origin, smoking status, alcohol consumption, SBP, DBP, HDLc, LDLc, anti-hypertension medication, lipid-lowering medication, history of diabetes, and eGFR.
**PICP:** C-terminal propeptide of procollagen type I; **hsTnT:** high sensitivity troponin T; **hsCRP:** high-sensitivity C reactive protein; **3-NT:** 3-nitrotyrosine; **NT-proBNP:** N-terminal propeptide of B-type natriuretic peptide.

At year 3 but not year 5, there was a smaller improvements and larger impairment in LAScd (-0.57, 95% CI: -1.10, -0.04) and LA PSLS (-0.82, 95% CI: -1.57, -0.06) among individuals with a higher baseline NT-proBNP after adjusting for all the covariates. At both years 3 and 5, LASi increased more among those who had a higher baseline NT-proBNP (year 3: 0.04, 95% CI 0.01, 0.06; year 5: 0.02, 95% CI 0.00, 0.04) in all the models. LAFi was observed to worsen faster with an elevated baseline PICP at year 3 after controlling for all the covariates (6.51, 95% CI: 0.26, 12.76). No other associations were found between baseline biomarkers and LA outcomes. (Table 3)

**Table 3.** Longitudinal association between baseline biomarkers and changes in left atrial structure and function at years 3 and 5, PREDIMED-Plus trial.

|  |  | PICP |  | hs-cTnT |  | CRP |  |
| --- | --- | --- | --- | --- | --- | --- | --- |
|  |  | Y3 vs. baseline | Y5 vs. baseline | Y3 vs. baseline | Y5 vs. baseline | Y3 vs. baseline | Y5 vs. baseline |
| **Left atrial conduit strain, %** |  |  |  |  |  |  |  |
|  | Crude | 0.28 (-0.81, 1.37) | 0.26 (-0.82, 1.34) | 0.99 (-0.01, 1.99) | 0.99 (0.00, 1.99) | -0.27 (-0.72, 0.19) | -0.40 (-0.87, 0.06) |
|  | Model 1 | 0.34 (-0.75, 1.42) | 0.34 (-0.74, 1.42) | 1.02 (0.02, 2.02) | 1.04 (0.04, 2.03) | -0.29 (-0.74, 0.17) | -0.41 (-0.87, 0.06) |
|  | Model 2 | 0.65 (-0.47, 1.76) | 0.42 (-0.71, 1.54) | 1.00 (-0.04, 2.04) | 0.67 (-0.37, 1.71) | -0.29 (-0.76, 0.17) | -0.40 (-0.88, 0.08) |
| **Left atrial contractile strain, %** |  |  |  |  |  |  |  |
|  | Crude | -0.05 (-1.19, 1.09) | -0.12 (-1.26, 1.01) | -0.06 (-1.11, 0.99) | -0.55 (-1.59, 0.49) | -0.31 (-0.79, 0.17) | 0.12 (-0.37, 0.61) |
|  | Model 1 | -0.03 (-1.17, 1.12) | -0.08 (-1.22, 1.05) | -0.04 (-1.09, 1.00) | -0.53 (-1.57, 0.51) | -0.32 (-0.79, 0.16) | 0.11 (-0.37, 0.60) |
|  | Model 2 | 0.04 (-1.14, 1.22) | -0.06 (-1.25, 1.13) | 0.00 (-1.10, 1.09) | -0.53 (-1.63, 0.56) | -0.26 (-0.75, 0.23) | 0.26 (-0.25, 0.76) |
| **Left atrial peak systolic longitudinal strain, %** |  |  |  |  |  |  |  |
|  | Crude | 0.39 (-1.18, 1.97) | 0.32 (-1.24, 1.89) | 1.08 (-0.37, 2.53) | 0.63 (-0.81, 2.07) | -0.53 (-1.19, 0.13) | -0.27 (-0.94, 0.40) |
|  | Model 1 | 0.44 (-1.13, 2.01) | 0.41 (-1.15, 1.97) | 1.10 (-0.35, 2.54) | 0.66 (-0.78, 2.10) | -0.54 (-1.21, 0.12) | -0.28 (-0.95, 0.40) |
|  | Model 2 | 0.73 (-0.87, 2.33) | 0.46 (-1.16, 2.08) | 1.07 (-0.42, 2.56) | 0.23 (-1.26, 1.73) | -0.51 (-1.17, 0.16) | -0.14 (-0.83, 0.55) |
| **Max left atrial volume, mL** |  |  |  |  |  |  |  |
|  | Crude | -1.61 (-4.70, 1.47) | -0.70 (-3.80, 2.40) | 1.05 (-1.78, 3.87) | 0.92 (-1.91, 3.75) | 0.22 (-1.08, 1.51) | -0.28 (-1.61, 1.05) |
|  | Model 1 | -1.50 (-4.58, 1.57) | -0.54 (-3.63, 2.55) | 1.07 (-1.75, 3.89) | 0.94 (-1.89, 3.77) | 0.21 (-1.08, 1.50) | -0.29 (-1.62, 1.03) |
|  | Model 2 | -1.00 (-4.16, 2.17) | -0.75 (-3.99, 2.50) | 1.36 (-1.57, 4.29) | 1.16 (-1.81, 4.13) | 0.05 (-1.26, 1.37) | -0.30 (-1.68, 1.08) |
| **Left atrial volume index, mL/m<sup>2</sup>** |  |  |  |  |  |  |  |
|  | Crude | -0.73 (-2.34, 0.87) | -0.16 (-1.77, 1.45) | 0.45 (-1.02, 1.92) | 0.32 (-1.16, 1.79) | 0.14 (-0.53, 0.81) | -0.13 (-0.82, 0.56) |
|  | Model 1 | -0.73 (-2.34, 0.87) | -0.16 (-1.77, 1.45) | 0.46 (-1.01, 1.92) | 0.32 (-1.16, 1.79) | 0.14 (-0.53, 0.81) | -0.13 (-0.82, 0.56) |
|  | Model 2 | -0.53 (-2.19, 1.13) | -0.49 (-2.18, 1.21) | 0.65 (-0.88, 2.19) | 0.40 (-1.16, 1.95) | 0.06 (-0.62, 0.75) | -0.16 (-0.88, 0.56) |
| **Left atrial ejection fraction, %** |  |  |  |  |  |  |  |
|  | Crude | 2.55 (0.19, 4.91) | 0.01 (-2.36, 2.38) | -0.53 (-2.73, 1.66) | -0.69 (-2.89, 1.51) | -0.53 (-1.53, 0.47) | 0.27 (-0.75, 1.30) |
|  | Model 1 | 2.65 (0.29, 5.00) | 0.16 (-2.21, 2.53) | -0.51 (-2.70, 1.68) | -0.66 (-2.85, 1.54) | -0.54 (-1.54, 0.46) | 0.27 (-0.76, 1.29) |
|  | Model 2 | 2.52 (0.12, 4.93) | 0.54 (-1.92, 3.00) | -0.26 (-2.52, 2.00) | -0.61 (-2.89, 1.68) | -0.43 (-1.44, 0.58) | 0.34 (-0.71, 1.40) |
| **Left atrial function index** |  |  |  |  |  |  |  |
|  | Crude | 6.28 (0.08, 12.49) | 2.14 (-4.06, 8.34) | -0.67 (-6.40, 5.07) | 1.60 (-4.08, 7.28) | -1.91 (-4.51, 0.69) | -1.17 (-3.83, 1.49) |
|  | Model 1 | 6.31 (0.11, 12.51) | 2.19 (-4.01, 8.39) | -0.68 (-6.41, 5.06) | 1.62 (-4.06, 7.30) | -1.93 (-4.53, 0.67) | -1.16 (-3.82, 1.50) |
|  | Model 2 | 6.51 (0.26, 12.76) | 3.72 (-2.67, 10.12) | -1.55 (-7.43, 4.33) | 1.18 (-4.68, 7.03) | -1.18 (-3.79, 1.44) | -0.61 (-3.33, 2.10) |
| **Left atrial stiffness index** |  |  |  |  |  |  |  |
|  | Crude | -0.02 (-0.07, 0.02) | 0.01 (-0.04, 0.06) | -0.02 (-0.06, 0.02) | 0.02 (-0.02, 0.06) | 0.01 (-0.01, 0.03) | 0.01 (-0.01, 0.03) |
|  | Model 1 | -0.02 (-0.07, 0.02) | 0.01 (-0.04, 0.05) | -0.02 (-0.06, 0.02) | 0.02 (-0.02, 0.06) | 0.01 (-0.01, 0.03) | 0.01 (-0.01, 0.03) |
|  | Model 2 | -0.02 (-0.07, 0.03) | 0.01 (-0.04, 0.06) | -0.02 (-0.06, 0.03) | 0.02 (-0.02, 0.07) | 0.01 (-0.01, 0.03) | 0.00 (-0.02, 0.03) |

**Table 3. Longitudinal association between baseline biomarkers and changes in left atrial structure and function at years 3 and 5, PREDIMED-Plus trial (Cont.)**
|  |  | 3-NT |  | NT-proBNP |  |
| --- | --- | --- | --- | --- | --- |
|  |  | Y3 vs. baseline | Y5 vs. baseline | Y3 vs. baseline | Y5 vs. baseline |
| Left atrial conduit strain, % |  |  |  |  |  |
|  | Crude | -0.36 (-0.84, 0.12) | -0.45 (-0.93, 0.04) | -0.55 (-1.07, -0.04) | -0.23 (-0.74, 0.28) |
|  | Model 1 | -0.36 (-0.84, 0.12) | -0.46 (-0.94, 0.02) | -0.58 (-1.09, -0.06) | -0.26 (-0.77, 0.25) |
|  | Model 2 | -0.42 (-0.91, 0.07) | -0.46 (-0.96, 0.05) | -0.57 (-1.10, -0.04) | -0.30 (-0.83, 0.24) |
| Left atrial contractile strain, % |  |  |  |  |  |
|  | Crude | 0.11 (-0.39, 0.61) | 0.03 (-0.47, 0.54) | -0.14 (-0.68, 0.40) | 0.37 (-0.17, 0.90) |
|  | Model 1 | 0.11 (-0.39, 0.61) | 0.03 (-0.48, 0.53) | -0.15 (-0.68, 0.39) | 0.36 (-0.18, 0.90) |
|  | Model 2 | 0.14 (-0.37, 0.65) | 0.08 (-0.45, 0.61) | -0.23 (-0.78, 0.33) | 0.28 (-0.28, 0.84) |
| Left atrial peak systolic longitudinal strain, % |  |  |  |  |  |
|  | Crude | -0.22 (-0.92, 0.48) | -0.39 (-1.09, 0.32) | -0.71 (-1.45, 0.04) | 0.14 (-0.60, 0.88) |
|  | Model 1 | -0.23 (-0.92, 0.47) | -0.40 (-1.10, 0.30) | -0.73 (-1.48, 0.01) | 0.11 (-0.63, 0.85) |
|  | Model 2 | -0.24 (-0.94, 0.46) | -0.34 (-1.06, 0.39) | -0.82 (-1.57, -0.06) | -0.01 (-0.77, 0.75) |
| Max left atrial volume, mL |  |  |  |  |  |
|  | Crude | 1.04 (-0.33, 2.40) | -0.87 (-2.25, 0.50) | 0.79 (-0.67, 2.25) | 0.12 (-1.34, 1.59) |
|  | Model 1 | 1.04 (-0.32, 2.41) | -0.86 (-2.23, 0.51) | 0.76 (-0.69, 2.22) | 0.08 (-1.38, 1.55) |
|  | Model 2 | 1.08 (-0.30, 2.47) | -1.17 (-2.60, 0.27) | 1.19 (-0.31, 2.69) | 0.42 (-1.11, 1.95) |
| Left atrial volume index, mL/m<sup>2</sup> |  |  |  |  |  |
|  | Crude | 0.50 (-0.20, 1.21) | -0.59 (-1.30, 0.12) | 0.42 (-0.34, 1.18) | 0.19 (-0.57, 0.95) |
|  | Model 1 | 0.50 (-0.20, 1.21) | -0.59 (-1.30, 0.13) | 0.42 (-0.34, 1.18) | 0.19 (-0.57, 0.95) |
|  | Model 2 | 0.60 (-0.12, 1.32) | -0.64 (-1.39, 0.11) | 0.72 (-0.06, 1.51) | 0.33 (-0.47, 1.12) |
| Left atrial ejection fraction, % |  |  |  |  |  |
|  | Crude | -0.76 (-1.82, 0.30) | -0.42 (-1.48, 0.65) | -0.50 (-1.63, 0.63) | -0.18 (-1.32, 0.95) |
|  | Model 1 | -0.76 (-1.82, 0.30) | -0.42 (-1.49, 0.65) | -0.52 (-1.65, 0.60) | -0.22 (-1.35, 0.91) |
|  | Model 2 | -0.83 (-1.90, 0.24) | -0.27 (-1.38, 0.84) | -0.64 (-1.79, 0.51) | -0.05 (-1.22, 1.12) |
| Left atrial function index |  |  |  |  |  |
|  | Crude | -2.02 (-4.77, 0.74) | 0.60 (-2.16, 3.36) | -1.39 (-4.33, 1.55) | 0.49 (-2.45, 3.43) |
|  | Model 1 | -2.02 (-4.77, 0.73) | 0.58 (-2.17, 3.34) | -1.39 (-4.33, 1.55) | 0.49 (-2.45, 3.43) |
|  | Model 2 | -2.57 (-5.33, 0.19) | 0.60 (-2.23, 3.44) | -2.52 (-5.50, 0.47) | -0.15 (-3.17, 2.88) |
| Left atrial stiffness index |  |  |  |  |  |
|  | Crude | 0.01 (-0.01, 0.03) | 0.00 (-0.02, 0.02) | 0.03 (0.01, 0.05) | 0.03 (0.01, 0.05) |
|  | Model 1 | 0.01 (-0.01, 0.03) | 0.00 (-0.02, 0.02) | 0.03 (0.01, 0.05) | 0.03 (0.01, 0.05) |
|  | Model 2 | 0.01 (-0.02, 0.03) | 0.00 (-0.03, 0.02) | 0.04 (0.01, 0.06) | 0.02 (0.00, 0.04) |

**Notes: Mixed models were used. The estimates correspond to 1 unit increase in log(biomarker concentration). Model 1: adjusted for age, sex; Model 2: further**
adjusted for further adjusted for BMI, origin, smoking status, alcohol consumption, SBP, DBP, HDLc, LDLc, anti-hypertension medication, lipid-lowering medication, history of diabetes, and eGFR.
PICP: C-terminal propeptide of procollagen type I; hsTnT: high sensitivity troponin T; hsCRP: high-sensitivity C reactive protein; 3-NT: 3-nitrotyrosine; NT-proBNP: N-terminal propeptide of B-type natriuretic peptide.

Throughout the 5-year follow-up period, a larger change in CRP was associated with a greater concomitant increase in LAScd (0.16, 95% CI: 0.05, 0.27) and a smaller decrease in LA PSLS (0.16, 95% CI: 0.01, 0.31). A higher increase in NT-proBNP was associated with a smaller enlargement in LASct (-0.18, 95% CI -0.30, -0.06), a larger decrease in LA PSLS (-0.19, 95% CI -0.35, -0.02) and LAEF (-0.34, 95% CI -0.59, -0.09), and increase in LAVmax (0.53, 95% CI 0.20, 0.86), and LAVi (0.28, 95% 0.10, 0.45). (Table 4) The model examining the association between changes in biomarker concentration at year 3 and changes in LA outcome from year 3 to year 5 revealed that a higher NT-proBNP change was related to a lower increase in LASct (-0.97, 95% CI -1.68, -0.26) and a higher increase in LASi (0.03, 95% CI 0.00, 0.05). (Table 5)

**Table 4.**
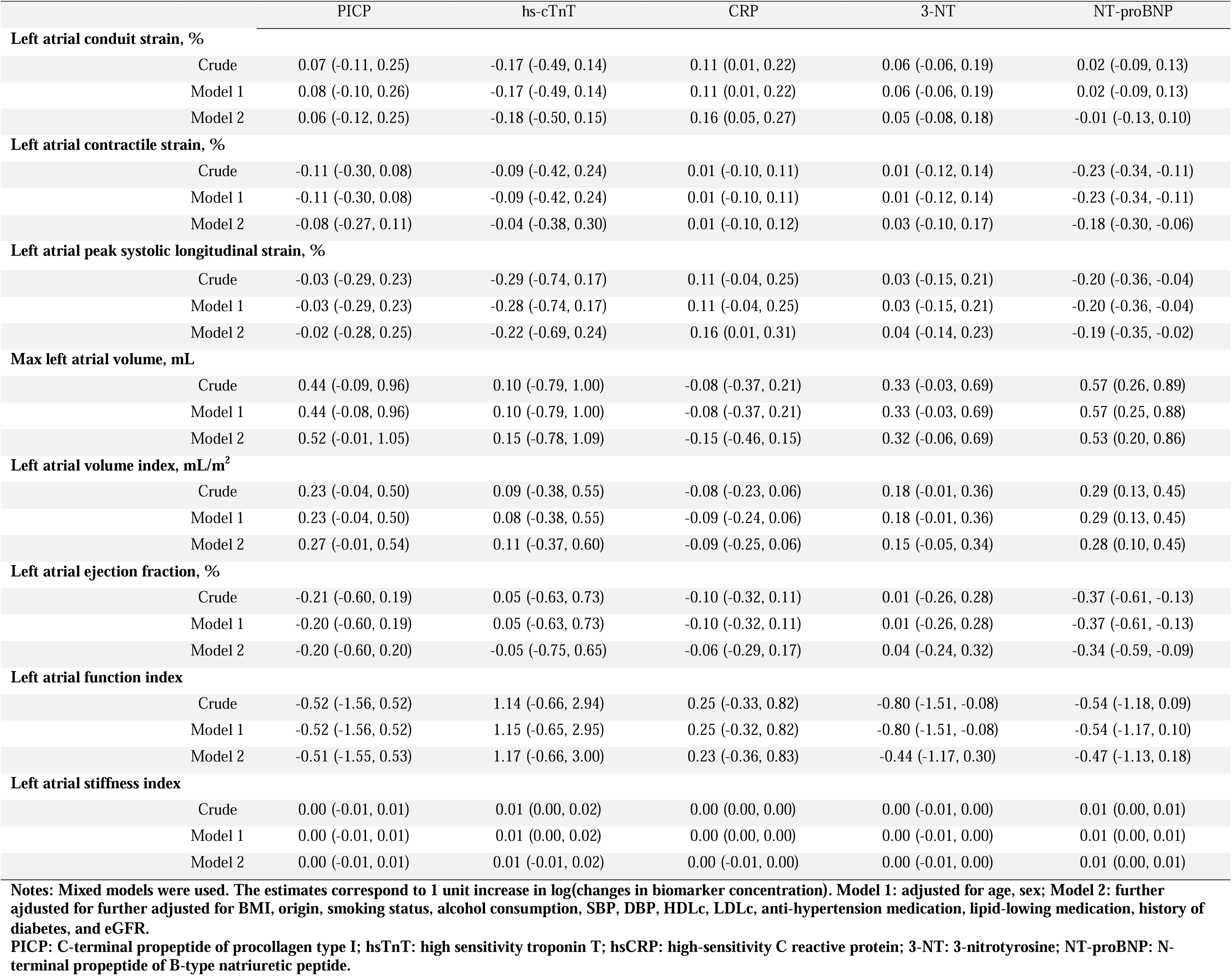
Associations between changes in biomarkers at year 5 and changes in left atrial structure and function at year 5, PREDIMED-Plus trial.

**Table 5.**
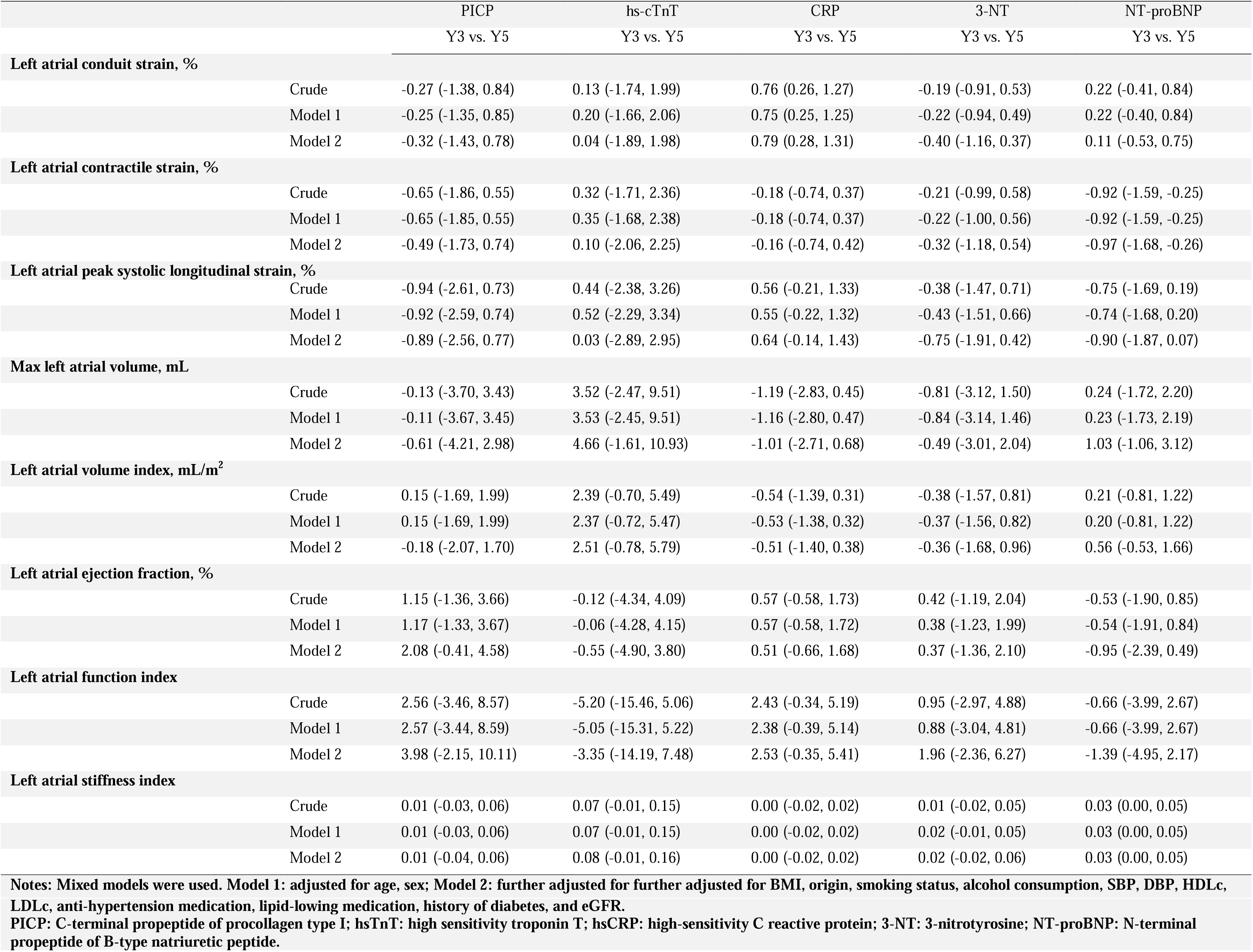
Longitudinal association between changes in biomarkers at year 3 and changes in left atrial structure and function at years 3 and 5, PREDIMED-Plus trial.

## Discussion

In this study of 532 adults with obesity or overweight in Spain, we found that higher NT-proBNP was cross-sectionally and longitudinally associated with enlarged LA volume, declining LA function, and elevated LA stiffness. The associations between NT-proBNP and worse LA strain were neither cross-sectionally nor longitudinally consistent regarding different LA strain measurements. Elevated hs-cTnT was observed to be related to higher LA volume and stiffness and impaired LA function cross-sectionally, but there were no similar results in other longitudinal models. PICP, CRP, and 3-NT did not show a significant effect on LA outcomes.

NT-proBNP is secreted from cardiac myocytes, mainly in a setting of myocyte stretch resulting from pressure overload and/or volume expansion.^23^ Existing literature suggests that NT-proBNP is a risk factor for heart failure and AF and can be clinically used for prediction purposes. ^15,24^ In the Multi-Ethnic Study of Atherosclerosis (MESA) cohort, findings showed that an increased NT-proBNP concentration over 10 years and high concentrations at both baseline and 10-year examinations were strongly associated with a larger increase in LA volume, greater decline in LA function and LA strain, compared to those with stable low concentrations.^25^ In that study, the study population was multiethnic and LA volumes were measured using cardiac magnetic resonance imaging (CMRI). Some evidence indicated that using echocardiography consistently underestimates LA volumes, compared to CMRI. Since our results are comparable with the findings in MESA, it suggests that our results may also reflect an underestimation. ^26,27^ In the Atherosclerosis Risk In Communities (ARIC) cohort study, the results suggested that NT-proBNP concentrations were robustly associated with LA structure and function measures cross-sectionally, including LAVmax, LAEF, LAScd, and LASct.^28^ Hs-TnT is a biomarker of myocardial injury which is released into the circulating blood after the injury occurs.^29^ Clinically, hs-TnT is used for acute coronary syndrome diagnosis.^29,30^ In a study that included 84 clinical patients in Germany, it was found that hs troponins (troponin T and I) were able to reflect LA function measured by CMRI. However, NT-proBNP did not show a significant impact on LA function.^31^ In a general multiethnic population in Dallas, both LAEF and LAmax were associated with NT-proBNP, and LAEF but not LAmax was found to be associated with hs-TnT cross-sectionally.^32^ To our knowledge, findings about the longitudinal associations between NT-proBNP and PICP and LA function and structure are limited. A few studies have reported how PICP and CRP were related to LA function and structure. Among patients with rheumatic heart disease, PICP was not significantly correlated with LAVi.^33^ CRP was unrelated to LAVmax or log-indexed LA volume longitudinally in an elderly population with high AF risk.^34^ Little is known about the impact of 3-NT on LA function and structure.

Our study findings are consistent with the prior evidence. However, further investigation is warranted regarding how the relationship between biomarkers and LA function and structure is involved in AF pathophysiology and development. Although the current analyses suggested a negative impact of higher NT-proBNP and possibly hs-TnT on LA impairment, the causal relationship cannot be inferred. Furthermore, given the concerns about cost and administration, it might be challenging to recommend routine TTE for those who are at high risk of AF. Our results may be useful for the design of therapeutic interventions, initiating routine TTE and AF risk assessment when abnormal elevation is noticed in blood biomarkers. Nevertheless, more evidence is needed to help with AF prevention and prediction.

Our study had several strengths. First, biomarker concentrations and LA outcomes were measured at 3 time points, at baseline and throughout the follow-up period, providing more information than single-time measurements. Additionally, the longitudinal analyses allowed us to glean insights into LA substrate development over time. Second, this ancillary study in PREDIMED-Plus trial had an excellent retention rate (above 96% at 2 years),^19^ a relatively large sample size compared to other studies, a core laboratory used for reading TTE studies, and great reproducibility. These advantages reduce the influence of the potential selection bias and information bias. Third, reliable and inclusive LA outcome measurements were obtained in the TTE studies. They enabled us to investigate biomarkers with multiple LA outcomes comprehensively. Our study also had some limitations. First, our study population is obese or overweight, at high risk of developing CVD, and primarily of European ancestry; therefore, the findings may not be generalizable to other populations. Second, the within-person and between-person variability and technical variability in the TTE studies may cause residual bias.

## Conclusion

In a population characterized by obesity or overweight with a high risk of CVD, elevated serum NT-proBNP was linked to the enlargement of LA volume and impairment in function over 5 years. Hs-TnT only demonstrated an association with LA volume enlargement and function impairment in a cross-sectional analysis. PICP, hsCRP, and 3-NT did not exhibit clear associations with LA function and structure. Further investigation is warranted for these associations in other populations to understand their implications for AF prevention.

## Disclosure

None.

## Data Availability

Data collaboration for PREDIMED-Plus study is guided by the Data Sharing and Management guide. We follow a controlled data collaboration model, using anonymised (de-identified) study data only, for collaborating with approved researchers. Requests are considered by the PREDIMED-Plus Steering Committee. Please see https://www.predimedplus.com/en/project/#top for detail.

## Acknowledgment

The authors would like to thank the colleagues, staff, and participants of the PREDIMED-Plus study for their important contributions.

